# Injury incidence, severity and type across the menstrual cycle in elite female professional footballers: a prospective three season cohort study

**DOI:** 10.1101/2023.07.12.23292497

**Authors:** Ally Barlow, Joanna M Blodgett, Sean Williams, Charles R Pedlar, Georgie Bruinvels

## Abstract

**Objectives:** The aim of the study was to assess the influence of menstrual cycle phase on injury incidence, severity and type in elite female professional footballers over three seasons.

**Methods:** Time-loss injuries and menstrual cycle data were prospectively recorded for 26 elite female football players across three seasons. The menstrual cycle was categorised into four phases using a standardised model: menstruation (phase 1; P1), remainder of follicular phase (phase 2; P2), early luteal (phase 3; P3), and pre-menstrual phase (phase 4; P4). Injury incidence rates (IRR) and ratios (IIRR) were calculated for overall injuries, injury type, contact vs non-contact, game/training and severity of injury.

**Results:** 593 cycles across 13,390 days were tracked during the study and 74 injuries from 26 players were eligible for analysis. Muscle injuries were the most prevalent sub-type (n=41). When comparing IRR between phases (reference: P1), injury rates were highest in P4 for overall (IIRR: 2.30 [95% CI: 0.99-5.34; p=0.05]), muscle-specific (6.07 [1.34-27.43; p=0.02]), non-contact (3.05 [1.10-8.50; p=0.03]) and ≤7 day’s time-loss injuries (4.40 [0.93-20.76; p=0.06]). Muscle-specific (IIRR P3:P1: 5.07 [1.16-22.07; p=0.03]) and ≤7 day’s time-loss (4.47 [1.01-19.68; p=0.05]) injury risk were also significantly higher in P3. No anterior cruciate ligament injuries were recorded across the monitoring period.

**Conclusion:** Injury risk was significantly elevated during the luteal phase of the menstrual cycle (P3 and P4) among elite female professional footballers. Further research is urgently needed to better understand the influence of the menstrual cycle on injury risk and to develop interventions to mitigate risk.

**What are the new findings?:**

- Injury risk was lowest in phase 1, during menstruation.
- Rate of overall injuries was highest in the luteal phase (phases 3 and 4), specifically for non-contact and lower severity injuries (e.g. 1-7 days missed training).
- Muscle injury risk increased across the menstrual cycle, with a greater incidence in the luteal phase (phases 3 and 4).
- Conversely to current perceptions, squad availability is significantly impacted by a wide range of injuries that are related to menstrual cycle phase, not just significant ligament injuries.

**How might it impact on clinical practice in the near future?:**

- When reviewing injury occurrence, mechanism and prevention guidelines, it is crucial to consider menstrual cycle day and phase.
- Longitudinal menstrual cycle monitoring provides an additional, potentially useful factor to support female athletes.
- The results of this study demonstrate that there is an urgent need to collect menstrual cycle data alongside standardised injury data to allow for further exploration in a larger sample.

## Introduction

Elite female football has recently seen a large increase in popularity and attention which has facilitated greater professionalisation of the game. This has resulted in an increased standard of play, more media attention and improved medical and performance support. Despite this, females are reported to have 21% more absence than males, primarily due to a greater incidence of severe ankle and knee ligament problems (1). For example, anterior cruciate ligament (ACL) injuries, are up to eight times more prevalent in females than male counterparts (1-3).

The menstrual cycle has been suggested as a factor which could modify injury risk in female athletes (4). Female sex hormones fluctuate throughout the menstrual cycle and can change by over 100% in 24-hours (5). These cyclical variations in endogenous hormones can have systemic effects, including influencing musculoskeletal tissue such as muscle, tendon, and bone (4, 6). Females must adjust and adapt to these hormonal changes, and this can affect readiness to play, with 51-93% of female athletes self-reporting performance detriments or negative experiences associated with the menstrual cycle (7, 8). Several studies have shown a greater incidence of anterior cruciate ligament (ACL) injury in the late follicular phase when oestrogen concentrations are at their peak (9-11). There are a number of potential hypotheses for this, including an increased ACL laxity in response to high levels of oestrogen (4). However, evidence is inconsistent, as some studies have also shown greater ACL injury incidence in the early follicular or late-luteal phases (12, 13).

To the knowledge of the authors, only two studies have examined all injury sub-types in women’s team sports alongside menstrual cycle data (14, 15), both of which considered a three phase approach and relied on inconsistent tracking (e.g. international camps spread over a four-year period), players to calculate menstrual phase or failed to exclude those on hormonal contraceptives. Research around the menstrual cycle has been limited by a lack of a standardised approach to prospective data collection and analysis. Some studies are confounded by the inclusion of combined hormonal contraceptive users, where exogenous hormones down regulate the natural hormonal (endogenous) rhythm (11-13, 16, 17). Another issue is that the definition of menstrual cycle phases varies between studies from the most simplistic (two phases) to the highly detailed (seven phases) (17). As highlighted above, previous studies have used three-phase models of the menstrual cycle including the menstrual phase (low oestrogen and progesterone; early follicular phase), pre-ovulation phase (high oestrogen; late follicular phase) and the luteal phase (high oestrogen and progesterone), failing to recognise the transitions between phases and the pre-menstrual phase. This approach does not account for the day-to-day variation in hormones, and ignores the fact that female athletes train and compete on any day of the menstrual cycle. These hormonal changes clearly have potential to affect injury risk and, therefore, warrant further investigation. Therefore, for the purpose of this study, the menstrual cycle will be divided into four phases: menstruation (early follicular), mid-late follicular, early to mid-luteal and pre-menstrual (late luteal).

To date, the incidence of injury, severity, and type in elite footballers has not been studied longitudinally alongside the menstrual cycle, despite the significant variations in hormonal concentrations, which have the potential to contribute towards injury risk. Therefore, the aim of this study is to investigate whether injury incidence, severity, and type significantly differ across the four phases of the menstrual cycle in a cohort of elite female footballers over three seasons.

## Methods

### Participants

Professional female footballers aged 18 and over, contracted to a single professional English Women’s Super League football club between June 2019 and May 2022 were invited to participate in the study. For inclusion in the study, players needed to have been eumenorrheic (10-16 periods per year for 12-months prior to the beginning of the study(18)). Players were excluded if they were using combined forms of hormonal contraception, pre-menarchal, oligomenorrheic or amenorrhoeic (absence of menstrual bleeding for > 90-days during the reproductive years (19)). Three players were excluded due to use of combined hormonal contraception, five players were excluded due to not providing consent and one player was excluded due to missing menstrual cycle data. No other players were excluded. In total, the present study included 26 participants and 593 tracked menstrual cycles. Prior to recruitment, players were provided with information about the research including terms of involvement. Informed consent was obtained before data analysis began. Participants were informed they could withdraw their data up to the date of data anonymisation without consequence. Ethical approval was granted by MSc SEM & SPY Research Ethics Approval Committee (at the University of Bath).

### Data Collection

This study utilised injury and menstrual cycle data that was prospectively collected over a three-year period. Injury data were collected by medical practitioners at the club and stored securely per European General Data Protection Regulation (GDPR) guidelines on the medical server at the club. Menstrual cycle data were collected using a mobile menstrual cycle tracking application, in accordance with GDPR guidelines (FitrWoman, Orreco, Ireland). A data sharing agreement between the club and the university was established to allow the data to be included in this research following individual player consent to inclusion.

### Injury recording and player availability

An injury was defined as an incident which prevented a player from taking part in full training or match-play for one or more days following the injury (Fuller 2006). Injuries sustained outside formal training or match-play were excluded from analysis. Injury information was recorded by the football club’s medical support staff and logged on an electronic medical records system (Kitman Labs, Dublin, Republic of Ireland). Each injury was classified using the Orchard Sport Injury Classification System (OSICS 10) (20). Injury severity was defined as the number of days of training and match play missed due to the injury sustained. Player availability status was recorded daily across the study period; one person-day indicated that an individual player participated in a training session or match.

### Menstrual Cycle Data

Players were screened on recruitment to establish whether they were using hormonal contraception or had any signs of menstrual dysfunction (e.g. amenorrhoea). Female health data were tracked longitudinally throughout the study, therefore any changes in menstrual status or use of hormonal contraception were carefully monitored. Throughout the study period, players tracked their own menstrual cycle data, recording menstruation days and intensity of flow. This information was accessible to medical support staff via a specific coaching platform. The mobile tracking application uses an integrated algorithm using a standardised model to divide the cycle into four phases based on an assumed hormonal profile. Briefly, total menstrual cycle length is used to retrospectively calculate phases 2 and 3, based on previous research evaluating and predicting the lengths of the follicular and luteal phases using thousands of menstrual cycles (21-23). Menstruation was classified as phase 1. The remainder of the follicular phase was classified as phase 2, phase 3 was the majority of the luteal phase, and phase 4 was the premenstrual window, defined as the 5 days prior to the onset of menstruation.

### Statistical Analysis

Descriptive statistics were used to describe the frequency of total injuries, injury type, and injury severity within each menstrual cycle phase. Given the differences in cycle length both within and between individuals, the frequency of injury was also visualised across cycle progression (where Day 1 of menstruation was considered 0% and the final day before menstruation was 100%), derived from the cycle length of the previous three full cycles. Person-days were quantified by summing the number of days of training/game participation from all individual players. This was calculated for each menstrual cycle phase and used to estimate injury incidence rate per 1,000 person-days for each phases(24). Injury incidence rate ratios were calculated by comparing injury incidence rates between phases, using phase 1 as the reference category. To assess statistical significance of the incidence rate ratios, we conducted Wald chi-square tests using an alpha of 0.05 (Rothman & Greenland, 2008). Analyses were carried out in Jamovi (V2.3) and Stata MP 17.0.

## Results

Player availability (e.g. non-rest days) and menstrual cycle data were recorded for a total of 13,390 days across the total study period; this included a total of 593 cycles tracked (mean cycle length:29±5 days) and an average of 239±41 days per player per season. During this time, 7,273 training and match person days were eligible for inclusion in analysis; illness, non-selection for matches and international break windows were not included for analysis. The average age of the 26 included players at baseline was 24.1±4.6 years.

A total of 74 eligible injuries occurred across the study period. The distribution of injury type is shown in Table 1; muscle injuries were the most common (n=41; 55.4%), followed by joint and ligament injuries (n=15; 20%). The majority of injuries were non-contact in nature (n=55; 74.3%), with more injuries occurring during games (n=42;56%) than training (n=33;44%). Injury severity ranged from 2 to 220 days with a median of 8 (Quartile 1:1.25, Quartile 3:14.75) days. The average cycle length when injury occurred was 29.9±4.6 days, while the average cycle length from the 3-months preceding injury was 29.5±3.4 days.

**Table 1:** Characteristics of Injuries (n=74) Collected Over 3-Seasons in Elite Female Football Players

| Injury summary (n=74) | Count (Median (IQR) or n (%) as appropriate) |
| --- | --- |
| Injury Type |  |
| - Muscle | 41 (55.4%) |
| - Joint and ligament | 15 (20%) |
| - Tendon | 7 (9.5%) |
| - Fracture & bone stress | 7 (9.5%) |
| - Central/Peripheral nervous system | 4 (5.4%) |
| Mechanism |  |
| - Non-contact | 55 (74.3%) |
| - Contact | 19 (25.7%) |
| Injury Occurrence |  |
| - Game | 42 (56%) |
| - Training | 33 (44%) |
| Injury severity (days) | 8 (1.25-14.75) |

IQR: interquartile range, SD: standard deviation.

Not standardising for phase length, more injuries occurred in phase 2 (n=26) and 3 (n=23) compared to phase 4 (n=17) and phase 1 (n=8). Figure 1A shows the distribution of injuries across each day of the cycle, while Figure 1B shows the distribution of injuries across cycle progression (i.e. as a proportion of each individual’s average cycle length). There was a slight increase in injuries before ovulation (∼20-40% into the cycle), with a more substantial peak towards the end of the cycle (∼70-90% into the cycle).

**Figure 1:**
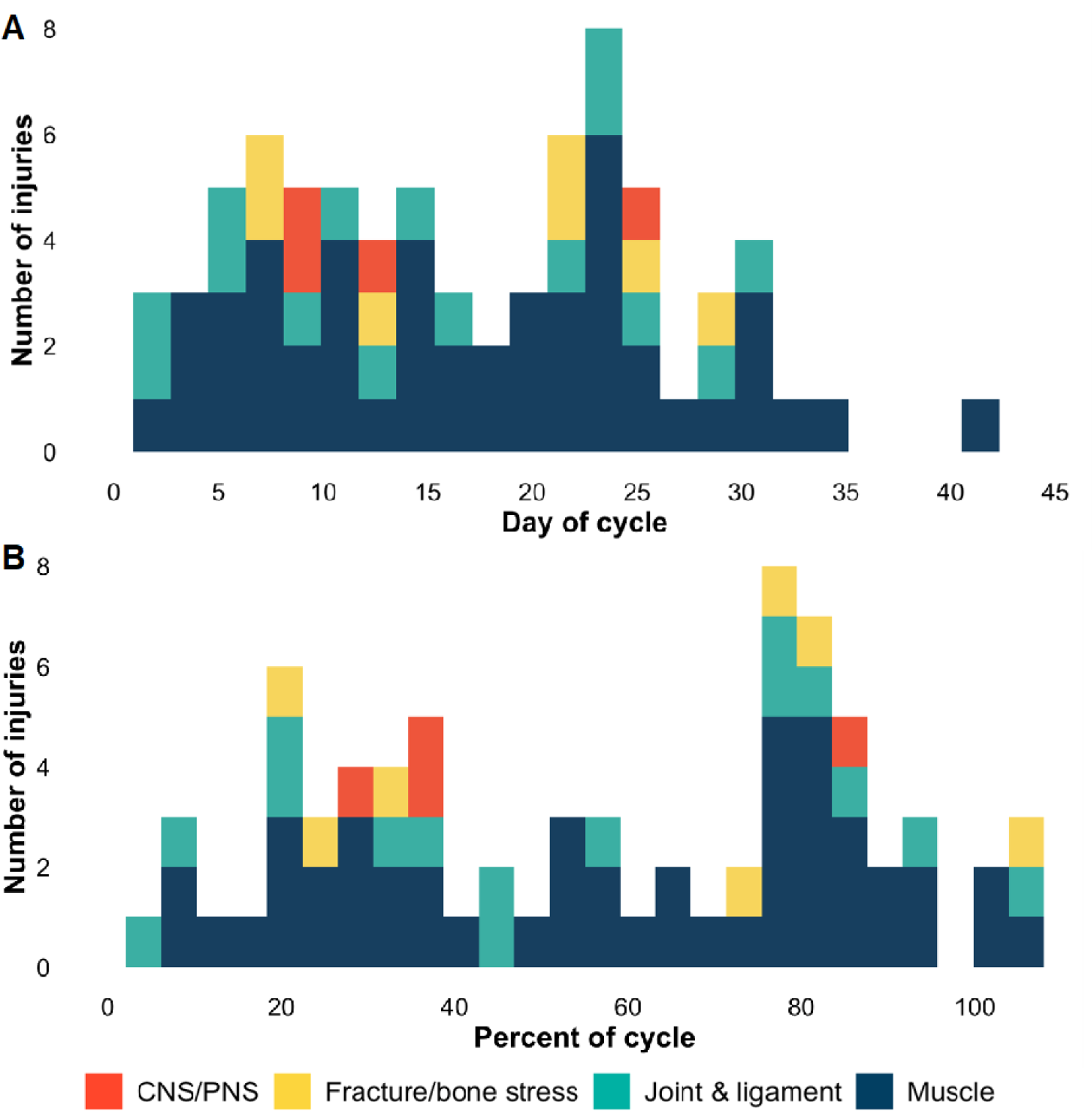
Stacked histogram demonstrating injury occurrence across A. menstrual cycle day (where 1=1^st^ day of menstruation) and B. menstrual cycle progression (where 0%= 1^st^ day of menstruation, 100% = final day before next menstruation).

Table 1 summarises the frequency and injury incidence rates across each menstrual cycle phase and provides incidence rate ratios comparing rates from phases 2,3 and 4 to phase 1. When using standardised incidence rates (per 1000 person days), injuries were least common in phase 1 (6.1 injuries per 1,000 person days; Table 2) and increased across the cycle (Phase 2: 9.9, Phase 3: 10.9, Phase 4: 14.0). Injury incidence rate ratios (IIRR) showed that injuries occurred 2.30 (95% CI: 0.99-5.34; p=0.05) times more frequently in phase 4 compared to phase 1. Although IIRR also suggested injury rates were higher in phase 3 (1.79; p=0.16) and phase 2 (1.62; p=0.23) compared to phase 1, this was not statistically significant.

**Table 2:** Total number of injuries, injury incidence rates, and injury incidence rate ratios across all injuries and by injury type.

|  | Total Number of Injuries |  |  |  |  | Injury Incidence Rate<br>(per 1000 person-days) |  |  |  | Injury Incidence Rate Ratios<br>(95% Confidence Interval, p-value)(ref: Phase 1) |  |  |
| --- | --- | --- | --- | --- | --- | --- | --- | --- | --- | --- | --- | --- |
|  | Total | Phase 1 | Phase 2 | Phase 3 | Phase 4 | Phase 1 | Phase 2 | Phase 3 | Phase 4 | Phase 2:1 | Phase 3:1 | Phase 4:1 |
| ALL INJURIES | 74 | 8 | 26 | 23 | 17 | 6.1 | 9.9 | 10.9 | 14.0 | 1.62<br>(0.73-3.59) p=0.23 | 1.79<br>(0.80-4.01) p=0.16 | 2.30<br>(0.99-5.34) p=0.05 |
| INJURY TYPE |  |  |  |  |  |  |  |  |  |  |  |  |
| Muscle | 41 | 2 | 12 | 16 | 11 | 1.5 | 4.6 | 7.6 | 9.1 | 3.07<br>(0.69-13.72) p=0.14 | 5.07<br>(1.16-22.07) p=0.03 | 6.07<br>(1.34-27.43) p=0.02 |
| Joint & Ligament | 15 | 3 | 6 | 3 | 3 | 2.3 | 2.2 | 1.4 | 2.5 | 0.96<br>(0.24-3.83) p=0.95 | 0.61<br>(0.12-3.02) p=0.54 | 1.09<br>(0.22-5.40) p=0.92 |
| Tendon | 7 | 3 | 2 | 1 | 1 | --<sup>a</sup> | --<sup>a</sup> | --<sup>a</sup> | --<sup>a</sup> | --<sup>a</sup> | --<sup>a</sup> | --<sup>a</sup> |
| Fracture & Bone Stress | 7 | 0 | 3 | 2 | 2 | --<sup>a</sup> | --<sup>a</sup> | --<sup>a</sup> | --<sup>a</sup> | --<sup>a</sup> | --<sup>a</sup> | --<sup>a</sup> |
| Central/Peripheral Nervous System | 4 | 0 | 3 | 1 | 0 | --<sup>a</sup> | --<sup>a</sup> | --<sup>a</sup> | --<sup>a</sup> | --<sup>a</sup> | --<sup>a</sup> | --<sup>a</sup> |
| CONTACT |  |  |  |  |  |  |  |  |  |  |  |  |
| Contact | 19 | 3 | 9 | 4 | 3 | 2.3 | 3.4 | 1.9 | 2.5 | 1.4<br>(0.40-5.47); p=0.56 | 0.83<br>(0.18-3.70) p=0.80 | 1.09<br>(0.22-5.40) p=0.92 |
| Non-contact | 55 | 5 | 17 | 19 | 14 | 3.8 | 6.5 | 9.0 | 11.6 | 1.71<br>(0.63-4.65) p=0.29 | 2.37<br>(0.88-6.36) p=0.09 | 3.05<br>(1.10-8.50) p=0.03 |
| SESSION TYPE |  |  |  |  |  |  |  |  |  |  |  |  |
| Game | 32 | 3 | 8 | 13 | 8 | 2.3 | 3.1 | 6.2 | 6.6 | 1.35<br>(0.36-5.09) p=0.66 | 2.70<br>(0.77-9.48) p=0.12 | 2.87<br>(0.76-10.84) p=0.12 |
| Training | 42 | 5 | 18 | 10 | 9 | 3.8 | 6.9 | 4.8 | 7.5 | 1.82<br>(0.67-4.90) p=0.24 | 1.26<br>(0.43-3.70) p=0.67 | 1.97<br>(0.66-5.91) p=0.22 |
| SEVERITY (days) |  |  |  |  |  |  |  |  |  |  |  |  |
| 1-7 | 35 | 2 | 11 | 14 | 8 | 1.5 | 4.2 | 6.7 | 6.6 | 2.80<br>(0.62-12.65) p=0.18 | 4.47<br>(1.01-19.68) p=0.05 | 4.40<br>(0.93-20.76) p=0.06 |
| 8-14 | 16 | 1 | 7 | 5 | 3 | 0.8 | 2.7 | 2.4 | 2.5 | 3.38<br>(0.41-27.46) p=0.26 | 3.00<br>(0.35-25.71) p=0.32 | 3.12<br>(0.32-30.08) p=0.32 |
| 15-28 | 11 | 3 | 3 | 3 | 2 | 2.3 | 1.1 | 1.4 | 1.7 | 0.48<br>(0.10-2.37) p=0.37 | 0.61<br>(0.12-3.02) p=0.54 | 0.74<br>(0.12-4.43) p=0.74 |
| >28 | 12 | 2 | 5 | 1 | 4 | 1.5 | 1.9 | 0.5 | 3.3 | 1.27<br>(0.25-6.54) p=0.78 | 0.33<br>(0.03-3.68) p=0.37 | 2.20<br>(0.40-12.03) p=0.36 |

<sup>a</sup> could not calculate due to insufficient injuries in these categories. Note: Confidence intervals are calculated in reference to Phase 1 differences

Further investigation by injury type suggested that muscle injuries were more common in phases 3 and 4 (7.6 and 9.1 injuries per 1000 person days) than in phases 1 and 2 (1.5 and 4.6 injuries per 1,000 person days). Using phase 1 as the reference, muscle injury rates were 3 times higher in phase 2 (3.07 [0.69-13.72]; p=0.14), 5.07 times higher in phase 3 (5.07 [1.16-22.07]; p=0.03), and 6.07 times higher in phase 4 (6.07 [1.34-27.43]; p=0.02). Injury incidence rates were similar for joint & ligament injuries across all phases (range:1.4 to 2.3 per 1,000 person days; all IIRR p>0.05), while ratios were not calculated for fracture and bone stress or central/peripheral nervous system sub-categories due to the limited number of the injuries of this type. Non-contact injuries were more prevalent (n=55) than contact injuries (n= 19); IIRR suggested that non-contact injury rates were higher in phase 4 compared to phase 1 (3.05 [1.10-8.50]; p=0.03), with no difference in contact injury rates across phases (all p>0.05). There was no indication that injury incidence rates differed across phases for more severe injuries (i.e. >7 days). However, when examining minimal/mild injury (i.e. 1-7 days lost), injury incidence rates were 4.47 (1.01-19.68; p=0.05) times higher in phase 3 and 4.40 (0.93-20.76) times higher in phase 4 compared to phase 1.

## Discussion

The present study assessed the influence of menstrual cycle phase on incidence, severity, and type of injury. Of the 74 injuries that occurred, muscle injuries were the most common, and injury incidence was greatest in phase 4 of the menstrual cycle (pre-menstrual phase), where 14 injuries occurred per 1,000 person days. When compared to phase 1, the incidence of muscle injuries was over three times more likely in phase 2, over five times more likely in phase 3, and over six times more likely in phase 4.

In contrast to our study, Lago-Fuentes et al. (15) used a three-phased menstrual cycle model including follicular, ovulatory and luteal phases to evaluate incidence of injury in elite female futsal players. When comparing injury incidence between phases, the highest incidence of injury occurred within the follicular phase, followed by the luteal phase, with lowest incidence in the ovulatory phase. Other studies have also observed greater injury incidence in the follicular phase, but these have only included ACL injuries (25, 26). Another recent study, applying a three phased model specifically looking at injuries in women’s football (14) found a higher incidence of injury, and specifically muscle and tendon injuries, within the late follicular phase. This finding is similar to Lago-Fuentes et. al., (15), both of which contradict findings from the present study.

The conflicting results from these studies when compared to the present study could be due to several factors. Given the significant hormonal changes that occur pre-menstrually, and the associated symptomology that occurs in this phase, we decided it more appropriate to separate the premenstrual phase from the rest of the luteal phase. Hormones vary substantially during the luteal phase and there is a high prevalence of symptoms specifically in the late luteal phase (7, 27). Another potential reason for the differences in results seen here when compared to previous research by Martin et al (14) and Lago-Fuentes (15) could be the ambiguity of menstrual cycle reporting, which was based on recall (14) or self-calculation (15). These approaches lead to more opportunity for error and potentially mis-classifying the phase of injury. Similarly to the present study, ovulation testing was not conducted, which we felt made it challenging to define an ovulation phase. A significant strength of the present study is that cycles were continuously tracked and phases were calculated used an established algorithm calculated through a menstrual health app, therefore, there was no recall bias in data collection.

The severity of ACL injuries has led to much research focus on this area; however, no ACL injuries were recorded in the present dataset during the three-year monitoring period. In fact, the present data highlights the need to consider other injury types, particularly muscle-based injuries given the patterns of results shown here. Relatively few joint and ligament injuries (n=15) were recorded in the current study, and these demonstrated an even distribution across the four phases (phase 1 = 2.3/1,000 person days, phase 2 = 2.3/1,000 person days, phase 3= 1.4 per 1,000 person days, phase 4 = 2.5/1,000 person days). However, more data are needed to better understand if there is an association between ligamentous injury and menstrual cycle phase; given the relative low prevalence of ACL injuries compared to muscle injuries, future study design must consider a larger sample size (e.g. several leagues rather than a single team).

There were more injuries observed in phases 3 and 4 in the present study (luteal phase). Specifically, the greatest incidence of injury was in phase 4. When considering injury type, muscle injuries showed the most variation across phases but were most common in phase 4. There are several potential reasons for the increased prevalence of muscle injuries here. Firstly, the higher levels of progesterone in the second half of the cycle (phases 3 and 4) have been associated with increased amino acid catabolism (29), thus increased muscle breakdown, which could potentially increase the risk of muscle injury during this phase. While it is important to highlight that we did not collect data to indicate an increase in progesterone in the luteal phase, the number of injuries collected should substantiate this point. Secondly, the withdrawal of hormones pre-menstrually, where there is a transition from high to low oestrogen and progesterone levels, results in the up-regulation of proinflammatory pathways and an increased expression of inflammatory mediators such as prostaglandins (30). The effects appear to be systemic, with a number of studies demonstrating elevated levels of inflammatory biomarkers peri-menstrually (31-33). This can lead to inflammation, which could affect recovery and induce a state of overload by compromising tissue integrity. Finally, the pre-menstrual window (phase 4) is where adverse menstrual cycle symptoms often occur; sleep disruption, altered mood, reduced co-ordination, and lower back pain are all frequently reported (7, 27), and these could potentially compromise readiness, movement patterns and recovery, also increasing injury risk. The hormonal changes are also likely to affect soft tissue action (4). Oestrogen increases muscle mass and strength, and the decline in oestrogen during phase 4 may be a factor in increasing muscle injury risk.

It is also notable that non-contact injuries were also three times more likely to occur in phase 4 versus phase 1. This could be associated with the effects of the hormonal withdrawal in phase 4. This is particularly notable as it is hypothesised that non-contact injuries are more preventable than contact injuries. Further research is needed to track inflammation, symptoms, and other factors of readiness such as sleep alongside the menstrual cycle phases. Building a multi-dimensional profile of female athletes from a medical, physiological, physical, and holistic perspective would facilitate a better understand of the potential injury considerations and lead to better player care.

While this data is novel and much needed, the findings here should be interpreted with caution, as a larger sample size is needed from multiple teams before making strong inferences to practice. However, this study indicates the potential value of monitoring the menstrual cycle and paves the way for future research to better identify how to support improvements in performance and reduce injury incidence in female athletes. The findings here suggest that it is crucial to collect continuous menstrual cycle data alongside standardised and comprehensive injury data. Notably, existing published guidelines for injury recording has no recommendation the inclusion of information on the menstrual cycle (34, 35). A consensus on how this information is collected needs to be established to allow large-scale collection and collaboration of useful and comparable data. Players need to be available to train and play on any given day in their cycle, therefore, pro-active management around the menstrual cycle such as cycle tracking, providing symptom support and education around nutrition, sleep and recovery should be considered on an individual basis, factoring in the presentation of their unique cycles. Future research should also consider ways this can be managed across the multi-disciplinary team. Despite much discussion focusing on ACL injuries in women’s sport, the findings here highlight that other injuries also need to be considered in female athletes. Given the association with muscle injuries and the menstrual cycle, future research should also capture self-reported symptoms alongside these as a potential means of risk mitigation.

### Limitations

There are some limitations which should be noted when interpreting the results of this study. The homogenous nature of the population, all being from a single club, is a clear strength of this study, however it may also lead to reduced generalisability as the included players were exposed to the same football environment, playing surfaces, training design and injury prevention protocols for the duration of their inclusion in this research.

Whilst the study was conducted over three seasons and includes a greater number of injuries and more person days than most previous studies published, the numbers remain relatively small, especially when categorising injuries into sub-types and studying incidence, severity, and type across 4 phases. Replicating this study design on a greater scale would require multiple teams to participate. Hormones were not tracked in this study. However, at present, the invasiveness and expense of collecting these measures longitudinally (i.e., over three seasons) is extremely challenging and impractical in an elite, high-performance environment. In time, as technology evolves to increase accuracy of low friction monitoring, research accuracy will significantly improve.

## Conclusions

This is the first study to prospectively investigate injury incidence, severity, and type across 4 phases of the menstrual cycle in elite female footballers. Overall injury incidence and specifically muscle injury incidence was highest in the pre-menstrual phase, with lowest risk observed during menstruation. This needs to be further investigated on a larger scale. Presently, with female participation in football and interest in the game globally at an all-time high the need for further understanding and investigation surrounding the impact of the menstrual cycle on injury is paramount (36). Elite female athletes need to be able to perform optimally on every day of their cycle so understanding the day-to-day variations of hormone cycles and their potential impact on injury and ways to mitigate this is necessary for practitioners to provide appropriate support.

## Data Availability

All data produced in the present study are available upon reasonable request to the authors

## EDI statement

Our research and author team included three female authors and two male authors, consisted of mid-career researchers and experienced senior researchers from different disciplines with three countries and two continents represented. The first and senior authors are both women.

Our study was on female football players. Our study population included females of different race, culture, ethnicity, socioeconomic level and included representation of marginalized groups such as women of colour, women from the LGBTQIA2S+ community and women from middle income countries.

## Acknowledgements

Thanks to the WSL Football club for supporting the study. Thanks to the medical team and players for their participation.

## Contributors

AB, GB and SW conceptualised the research study aims and methodology; AB, GB, and JB curated, reviewed and categorised all data provided by the football club; AB and JB conducted the formal statistical analysis; AB, GB, JB and CP wrote the manuscript; All authors reviewed and approved the final manuscript.

## Data sharing statement

The data included in this study will not be made publicly available to protect athlete confidentiality.

## Patient and public involvement

Patients or members of the public were not involved in the design, or conduct, or reporting, or dissemination plans of our research.

